# Predictors of second COVID-19 booster dose or new COVID-19 vaccine hesitancy among nurses: a cross-sectional study

**DOI:** 10.1101/2022.06.04.22275989

**Authors:** Petros Galanis, Irene Vraka, Aglaia Katsiroumpa, Olga Siskou, Olympia Konstantakopoulou, Theodoros Katsoulas, Theodoros Mariolis-Sapsakos, Daphne Kaitelidou

**Author notes:** Corresponding author: Petros Galanis, Assistant Professor, Clinical Epidemiology Laboratory, Faculty of Nursing, National and Kapodistrian University of Athens, 123 Papadiamantopoulou street, GR-11527, Athens, Greece. Funding: None. Declarations of interest: None.

## Abstract

**Aims and objectives:** To assess the levels of second COVID-19 booster dose or new COVID-19 vaccine hesitancy among nurses and explore the potential predictors of vaccine hesitancy.

**Background:** COVID-19 full vaccination seems to be highly effective against highly contagious variants of SARS-CoV-2. Healthcare workers are a high-risk group since they have experienced high levels of COVID-19-associated morbidity and mortality.

**Methods:** An on-line cross-sectional study was carried out in Greece in May 2022, using a self-administered questionnaire. The study population included nurses in healthcare services who were fully vaccinated against COVID-19 at the time of study. We considered socio-demographic characteristics, COVID-19-related variables, and attitudes toward COVID-19 vaccination and pandemic as potential predictors of vaccine hesitancy.

**Results:** Among 795 nurses, 30.9% were hesitant toward a second booster dose or a new COVID-19 vaccine. Independent predictors of hesitancy included lower educational level, absence of a chronic condition, good/very good self-perceived physical health, lack of flu vaccination during 2021, front-line nurses that provided healthcare to COVID-19 patients during the pandemic, nurses that had not been diagnosed with COVID-19 during the pandemic, and nurses that had at least one relative/friend that has died from COVID-19. Moreover, increased compliance with hygiene measures, increased fear of a second booster dose/new COVID-19 vaccine, and decreased trust in COVID-19 vaccination were associated with increased hesitancy.

**Conclusions:** Our study shows that a significant percentage of nurses are hesitant toward a second booster dose/new COVID-19 vaccine. This initial hesitancy could be a barrier to efforts to control the COVID-19 pandemic. There is a need to communicate COVID-19 vaccine science in a way that is accessible to nurses in order to decrease COVID-19 vaccine hesitancy.

## 1. Introduction

COVID-19 vaccines efficacy or effectiveness against severe disease remains high but decreases somewhat within six months after the primer doses (Feikin et al., 2022; Iheanacho et al., 2021). However, decrease in efficacy or effectiveness against infection and symptomatic disease is higher (Feikin et al., 2022). Moreover, the emergence of various highly contagious variants of SARS-CoV-2 compromises more the effectiveness of COVID-19 vaccines. Full vaccination seems to be highly effective against Alpha variant, but moderate effective against Beta, Gamma, and Delta variants (Zeng et al., 2022).

## 2. Background

Several studies suggest that administration of a COVID-19 booster dose improves the immunogenicity of the vaccine and prolongs protection (Bar-On et al., 2021; Falsey et al., 2021; Jantarabenjakul et al., 2022). Early evidence from a study in Israel shows high vaccine immunoreactivity in healthcare workers (Saiag et al., 2021). Moreover, the rates of confirmed COVID-19 cases, admission to hospital, COVID-19-related death, and severity of disease are lower among those who received a booster dose (Barda et al., 2021; Kamar et al., 2021). However, waning immunity to a first booster and evolving highly contagious variants of SARS-CoV-2 have led to the consensus recommendation of prioritizing high-risk groups (e.g. oncologic patients, HIV patients, organ transplant recipients, and adults 65 years of age or older with comorbidities) for a second booster dose in various countries (Jantarabenjakul et al., 2022; Patalon et al., 2022).

Healthcare workers are a high-risk group since they have experienced high levels of COVID-19-associated morbidity and mortality (International Council of Nurses., 2022; World Health Organization., 2021). Thus, the National Nurses United, the largest union and professional association of registered nurses in the USA, has already called the Centers for Disease Control to approve a recommendation for a second booster dose for nurses (National Nurses United., n.d.). Moreover, the Joint Committee on Vaccination and Immunisation in the United Kingdom advises a second booster for vulnerable adults and healthcare workers in autumn 2022 in order to increase immunity in high-risk groups and to decrease admissions to hospital and COVID-19-related deaths, over the winter period (Wise, 2022). Thus, recommendation of a second COVID-19 booster dose or a new COVID-19 vaccine for healthcare workers in autumn 2022 seems to be a reasonable scenario.

Hesitancy of healthcare workers to accept a second booster/new COVID-19 vaccine could be a significant public health treat with negative effects on individuals and their communities. Until now, levels of second booster/ new COVID-19 vaccine hesitancy among healthcare workers are not investigated. Moreover, only a few studies have estimated the vaccine hesitancy towards the first booster dose among healthcare workers (Alhasan et al., 2021; Chrissian et al., 2022; Klugar et al., 2021; Koh et al., 2022). Also, literature is scarce regarding the factors that affect healthcare workers decision to accept the first COVID-19 booster dose (Alhasan et al., 2021; Chrissian et al., 2022; Klugar et al., 2021). Vaccine hesitancy is higher among females, younger healthcare workers, those with a lower educational level, and those with low levels of knowledge and confidence in COVID-19 booster doses.

Therefore, this study aimed to assess the levels of second COVID-19 booster dose or new COVID-19 vaccine hesitancy among nurses and explore the potential predictors of vaccine hesitancy.

## 3. Methods

### 3.1. Study design and participants

A cross-sectional study with a convenience sample was carried out in Greece between 23 and 30 May 2022, using a self-administered questionnaire. At the time of data collection, the Greek Committee on Vaccination had recommended a second booster dose only to vulnerable fully vaccinated groups, e.g. adults 70 years of age or older, adults 60 years of age or older with comorbidities, and older persons in nursing or care homes. Vulnerable individuals that they received a first booster dose at least four months ago were eligible to receive a second booster. Thus, at the time of the study, a second booster was not recommended to nurses. The inclusion criteria for the study were nurses in healthcare services (e.g. hospital, health centers, nursing homes, etc.) who have completed a COVID-19 vaccine course (e.g. primer doses of a two-dose COVID-19 vaccine and a first booster dose). We created an anonymous version of the study questionnaire with Google forms and we used social media platforms to distribute it. Also, study investigators sent the questionnaire via e-mail to all their contacts that are nurses in healthcare services. Moreover, we asked the participants to send the questionnaire to nurses they know. Thus, a snowball sampling technique was applied. We applied the STROBE checklist in our study.

### 3.2. Predictor variables

We considered socio-demographic characteristics, COVID-19 related variables, and attitudes toward COVID-19 vaccination and pandemic as potential predictors of vaccine hesitancy among nurses.

The first part of the study questionnaire included the following socio-demographic characteristics: gender (female or male), age (continuous variable), marital status (single, married, divorced, or widow), MSc or/and PhD degree (no or yes), chronic disease (no or yes), self-assessment of physical health (very poor, poor, average, good, or very good), influenza vaccination during 2021 (no or yes), front-line nurses that provided healthcare services to COVID-19 patients during the pandemic (no or yes), and years of clinical experience (continuous variable).

The second part of the questionnaire included COVID-19-related variables, such as previous COVID-19 diagnosis (no or yes), COVID-19-related death in family members/friends (no or yes), and adverse reactions and discomfort experienced after previous COVID-19 vaccine doses (scale from 0 [none] to 10 [great discomfort]).

The third part of the questionnaire included variables regarding nurses’ attitudes toward COVID-19 vaccination and pandemic. We used a 16-items valid questionnaire in Greek population to measure attitudes toward COVID-19 vaccination and pandemic (Galanis et al., 2022). This questionnaire measures fear of COVID-19, information regarding COVID-19 pandemic and vaccination, compliance with hygiene measures, and trust in COVID-19 vaccination. We calculated a total score from 0 to 10 for each factor with higher values indicating higher level of fear, information, compliance and trust. Internal reliability for the questionnaire was very good in our study; Cronbach’s coefficient alpha was 0.87, 0.77, 0.86, and 0.76 for the four factors respectively.

Finally, nurses’ attitudes toward a second booster dose or a new COVID-19 vaccine were measured with three study developed items; “I am afraid to have a second booster dose or a new COVID-19 vaccine”, “I worry about the long-term side effects of a second booster dose or a new COVID-19 vaccine”, and “I feel protected by the previous COVID-19 vaccine doses”. Response options ranging from 0 (totally disagree) to 10 (totally agree).

### 3.3. Outcome variable

Pharmaceutical companies seek now for the next generation of COVID-19 vaccines conferring longer duration of protection and increasing immunity against SARS-CoV-2 variants (Nohynek & Wilder-Smith, 2022). Thus, we investigated nurses’ hesitancy toward a second booster dose or a new COVID-19 vaccine since a new vaccine in fall of 2022 seems to be possible. Nurses’ hesitancy toward a second booster dose or a new COVID-19 vaccine was measured with a study developed item; “If a second booster dose or a new COVID-19 vaccine is recommended as a supplement to the current vaccination schedule, would you accept it?”. World Health Organization defines vaccine hesitancy as the delay in acceptance or refusal of vaccination despite availability (MacDonald, 2015). Therefore, we considered the answers “definitely no”, “probably no”, and “unsure” indicative of future booster/vaccine hesitancy, and the answers “probably yes”, and “definitely yes” indicative of future booster/vaccine acceptance. Thus, we handled the vaccine hesitancy as a dichotomous variable in the statistical analysis.

Also, we asked nurses to select the reasons for their hesitancy including concerns about COVID-19 vaccines safety and effectiveness, short-term and long-term side effects, COVID-19-related complications, tiredness due to the vaccination procedure, effective immunization against the SARS-CoV-2, and necessity of a second booster dose or a new COVID-19 vaccine.

### 3.4. Sample size calculation

Since there are no studies regarding nurses’ hesitancy toward a second booster dose or a new COVID-19 vaccine, we based on studies that estimate healthcare workers hesitancy toward the first booster in order to find out the minimum sample size (Alhasan et al., 2021; Chrissian et al., 2022; Klugar et al., 2021; Koh et al., 2022). These studies found that median healthcare workers hesitancy toward the first booster was 31%. Considering a similar hesitancy for a second booster/new vaccine, a confidence level of 95%, and a margin of error 4%, a minimum sample size of 514 nurses was obtained. We decided to recruit a larger sample (n=795) in order to decrease random error. Performing a sensitivity analysis with different vaccine hesitancy rates, our sample exceeded the minimum sample size; minimum sample size of 385 nurses for a vaccine hesitancy rate of 20%, and minimum sample size of 505 nurses for a vaccine hesitancy rate of 70%.

### 3.5. Ethical issues

Participation in our study was anonymous and voluntary. We informed the participants about the aim of the study and the survey questionnaire. The Ethics Committee of Department of Nursing, National and Kapodistrian University of Athens approved the study protocol (reference number; 370, 02-09-2021). Moreover, we applied the principles of the Declaration of Helsinki.

### 3.6. Statistical analysis

We use frequencies and percentages to describe categorical variables, while we use mean and standard deviation to describe continuous variables. As explained above we handled the vaccine hesitancy as a dichotomous variable in the statistical analysis. Thus, we used univariate and multivariable logistic regression analysis to assess the associations between predictor variables and vaccine hesitancy. We calculated unadjusted and adjusted odds ratios (aOR), 95% confidence intervals (CI) and p-values. The alpha significance level was set at the level of 0.05. Statistical analysis was performed with the Statistical Package for Social Sciences software (IBM Corp. Released 2012. IBM SPSS Statistics for Windows, Version 21.0. Armonk, NY: IBM Corp.).

## 4. Results

### 4.1. Socio-demographic characteristics

A total of 795 nurses completed the study questionnaire. The majority of nurses was females (81.8%) and married (56%). The mean age of the nurses was 38.5 years old. At least one chronic disease was present in 20.8% of the nurses, while 89.3% reported that their physical health was good/very good. Nearly half of the nurses (50.3%) received a flu vaccine during 2021. Among nurses, 53.5% provided healthcare services to COVID-19 patients during the pandemic. The mean number of years of clinical experience was 14.8. Table 1 presents the socio-demographic characteristics of the nurses.

**Table 1.** Socio-demographic characteristics of the 795 nurses.

| **Characteristics** | **N** | **%** |
| --- | --- | --- |
| Gender |  |  |
| Males | 145 | 18.2 |
| Females | 650 | 81.8 |
| Age (years) <sup>†</sup> | 38.5 | 10.6 |
| Marital status |  |  |
| Singles | 325 | 40.9 |
| Married | 445 | 56.0 |
| Divorced | 20 | 2.5 |
| Widowed | 5 | 0.6 |
| MSc or/and PhD degree |  |  |
| No | 210 | 26.4 |
| Yes | 585 | 73.6 |
| Chronic disease |  |  |
| No | 630 | 79.2 |
| Yes | 165 | 20.8 |
| Self-perceived physical health |  |  |
| Very poor | 5 | 0.6 |
| Poor | 5 | 0.6 |
| Moderate | 75 | 9.4 |
| Good | 340 | 42.8 |
| Very good | 370 | 46.5 |
| Influenza vaccination during 2021 |  |  |
| No | 395 | 49.7 |
| Yes | 400 | 50.3 |
| Front-line nurses |  |  |
| No | 370 | 46.5 |
| Yes | 425 | 53.5 |
| Years of clinical experience<sup>a</sup> | 14.8 | 10.8 |

<sup>†</sup> mean, standard deviation

### 4.2. Hesitancy and attitudes toward second booster dose/new COVID-19 vaccine

Nurses’ hesitancy and attitudes toward a second booster dose or a new COVID-19 vaccine are shown in Table 2. Of the entire sample, 245 (30.9%) were hesitant toward a second booster dose or a new COVID-19 vaccine. In particular, 15.1% were unsure, 10.1% had not definitively decided that they would not take a second booster dose/new COVID-19 vaccine, and 5.7% had definitively decided that they would not take a second booster dose/new COVID-19 vaccine. On the other hand, 37.7% of the nurses had definitively decided that they would take a second booster dose/new COVID-19 vaccine and 31.4% had not definitively decided that they would take a second booster dose/new COVID-19 vaccine.

**Table 2.**
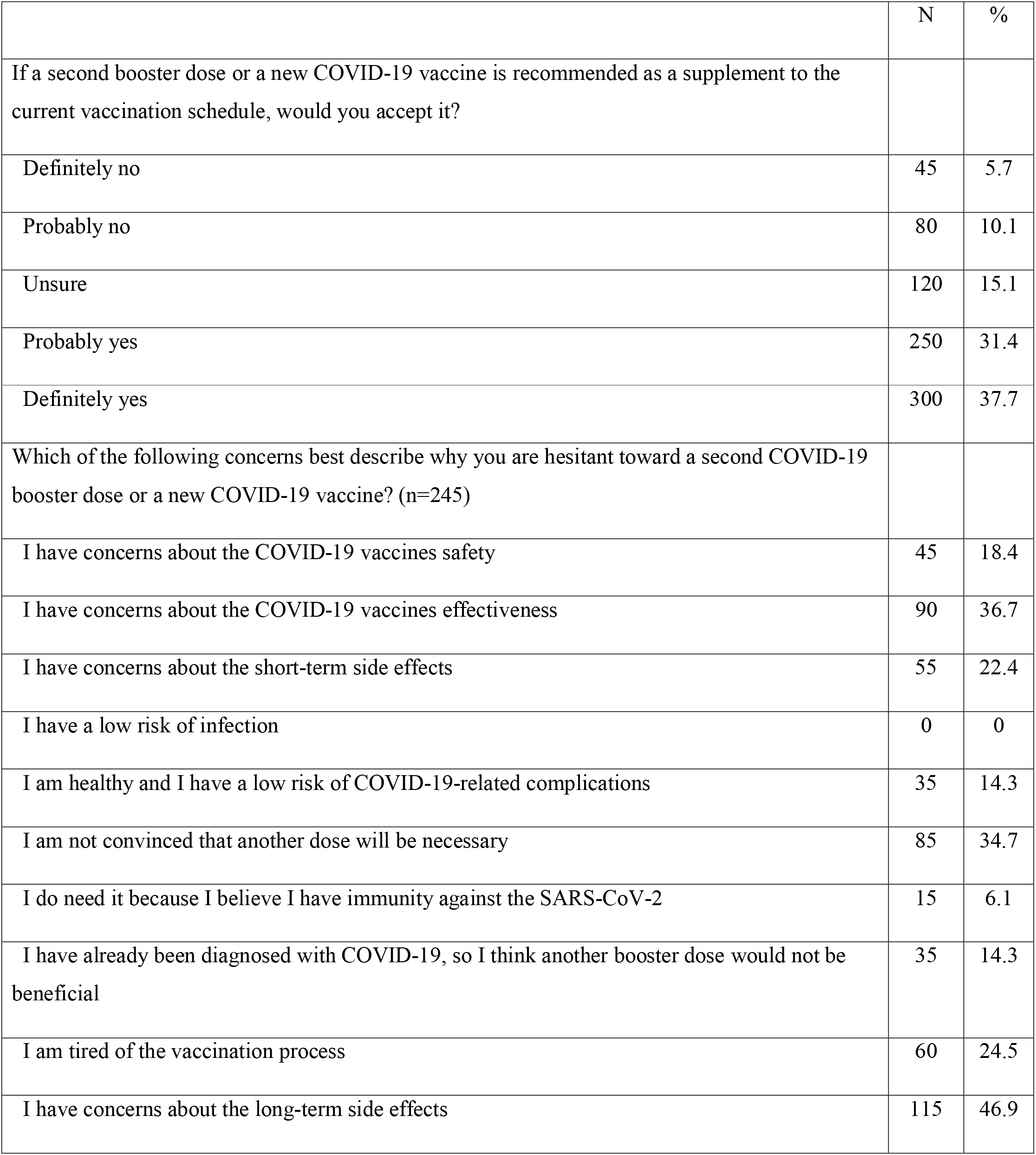
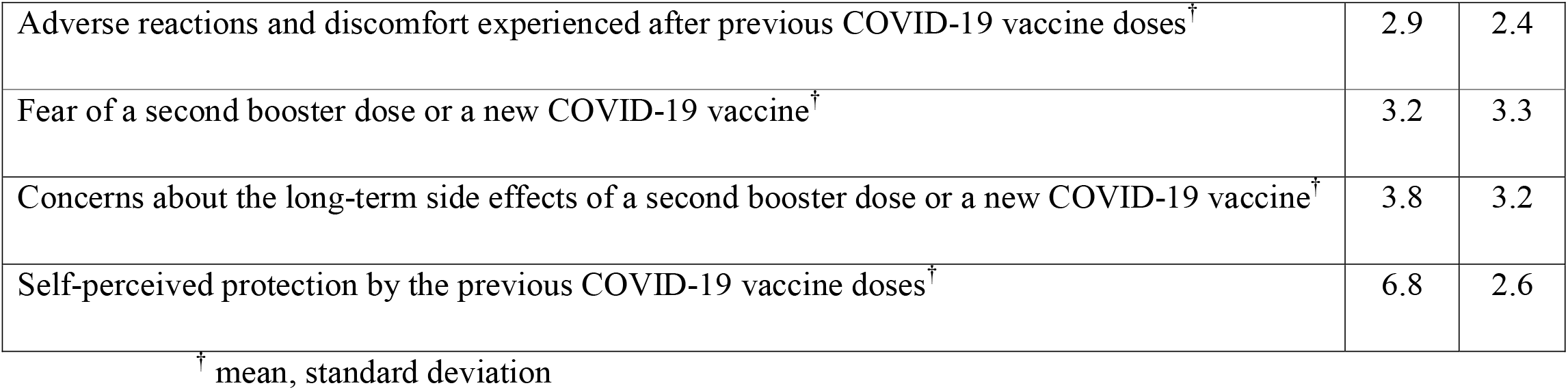
Nurses’ hesitancy and attitudes toward a second booster dose or a new COVID-19 vaccine (n=795).

|  | N | % |
| --- | --- | --- |
| If a second booster dose or a new COVID-19 vaccine is recommended as a supplement to the current vaccination schedule, would you accept it? |  |  |
| Definitely no | 45 | 5.7 |
| Probably no | 80 | 10.1 |
| Unsure | 120 | 15.1 |
| Probably yes | 250 | 31.4 |
| Definitely yes | 300 | 37.7 |
| Which of the following concerns best describe why you are hesitant toward a second COVID-19 booster dose or a new COVID-19 vaccine? (n=245) |  |  |
| I have concerns about the COVID-19 vaccines safety | 45 | 18.4 |
| I have concerns about the COVID-19 vaccines effectiveness | 90 | 36.7 |
| I have concerns about the short-term side effects | 55 | 22.4 |
| I have a low risk of infection | 0 | 0 |
| I am healthy and I have a low risk of COVID-19-related complications | 35 | 14.3 |
| I am not convinced that another dose will be necessary | 85 | 34.7 |
| I do need it because I believe I have immunity against the SARS-CoV-2 | 15 | 6.1 |
| I have already been diagnosed with COVID-19, so I think another booster dose would not be beneficial | 35 | 14.3 |
| I am tired of the vaccination process | 60 | 24.5 |
| I have concerns about the long-term side effects | 115 | 46.9 |
| Adverse reactions and discomfort experienced after previous COVID-19 vaccine doses<sup>†</sup> | 2.9 | 2.4 |
| Fear of a second booster dose or a new COVID-19 vaccine<sup>†</sup> | 3.2 | 3.3 |
| Concerns about the long-term side effects of a second booster dose or a new COVID-19 vaccine<sup>†</sup> | 3.8 | 3.2 |
| Self-perceived protection by the previous COVID-19 vaccine doses<sup>†</sup> | 6.8 | 2.6 |

<sup>†</sup> mean, standard deviation

When nurses asked about the reasons for not receiving a second booster dose/new COVID-19 vaccine, the most common reasons were concerns about the long-term side effects (46.9%), concerns about the COVID-19 vaccines effectiveness (36.7%), non-necessity of a second booster dose/new COVID-19 vaccine (34.7%), tiredness due to the vaccination procedure (24.5%), concerns about the short-term side effects (22.4%), and concerns about the COVID-19 vaccines safety (18.4%).

Adverse reactions and discomfort experienced after previous COVID-19 vaccine doses were low (mean value of 2.9 in a scale from 0 to 10). Moreover, nurses reported low levels of fear of a second booster dose/new COVID-19 vaccine (mean value of 3.2 in a scale from 0 to 10), but moderate level of concerns about the long-term side effects (mean value of 3.8 in a scale from 0 to 10).

### 4.3. COVID-19-related variables and attitudes toward COVID-19 vaccination and pandemic

More than half of the nurses (56.6%) had been diagnosed with COVID-19. Almost one out of four nurses (23.9%) had at least one relative/friend that has died from COVID-19. Nurses reported moderate levels of fear of COVID-19 (mean value of 5.8 in a scale from 0 to 10) and moderate to high levels of trust in COVID-19 vaccination (mean value of 7.1 in a scale from 0 to 10). Moreover, nurses were highly compliant with the hygiene measures (mean value of 9.5 in a scale from 0 to 10) and they believed that they are well-informed regarding the COVID-19 pandemic and vaccination (mean value of 8.7 in a scale from 0 to 10). Table 3 depicts COVID-19-related variables and nurses’ attitudes toward COVID-19 vaccination and pandemic.

**Table 3.** COVID-19-related variables and nurses’ attitudes toward COVID-19 vaccination and pandemic (n=795).

|  | N | % |
| --- | --- | --- |
| Previous COVID-19 diagnosis |  |  |
| No | 345 | 43.4 |
| Yes | 450 | 56.6 |
| COVID-19-related death in family members/friends |  |  |
| No | 605 | 76.1 |
| Yes | 190 | 23.9 |
| Fear of COVID-19<sup>†</sup> | 5.8 | 2.2 |
| Information regarding the COVID-19 pandemic and vaccination<sup>†</sup> | 8.7 | 1.4 |
| Compliance with hygiene measures<sup>†</sup> | 9.5 | 0.8 |
| Trust in COVID-19 vaccination<sup>†</sup> | 7.1 | 1.7 |

<sup>†</sup> mean, standard deviation

### 4.4. Predictors of second booster dose/new COVID-19 vaccine hesitancy

Independent predictors of hesitancy toward a second booster dose/new COVID-19 vaccine included the following socio-demographic characteristics: lower educational level (aOR = 9.16, 95% CI: 4.08-20.57), married status (aOR = 3.20, 95% CI: 1.35-7.56), absence of a chronic disease (aOR = 3.19, 95% CI: 1.29-7.93), good/very good self-perceived physical health (aOR = 8.05, 95% CI: 2.63-24.65), lack of flu vaccination during 2021 (aOR = 9.40, 95% CI: 4.40-20.09), and front-line nurses that provided healthcare services to COVID-19 patients during the pandemic (aOR = 7.85, 95% CI: 3.23-19.05). Moreover, nurses that had not been diagnosed with COVID-19 during the pandemic (aOR = 5.59, 95% CI: 2.50-12.51) and those that had at least one relative/friend that has died from COVID-19 (aOR = 6.77, 95% CI: 2.75-16.65) were more likely to be hesitant. In addition, increased compliance with hygiene measures (aOR = 1.75, 95% CI: 1.18-2.59), increased fear of a second booster dose or a new COVID-19 vaccine (aOR = 1.73, 95% CI: 1.50-1.99), and decreased trust in COVID-19 vaccination (aOR = 0.27, 95% CI: 0.19-0.38) were associated with increased hesitancy. Detailed results from univariate and multivariable logistic regression analysis with second booster dose/new COVID-19 vaccine hesitancy as the dependent variable are shown in Table 4.

**Table 4.**
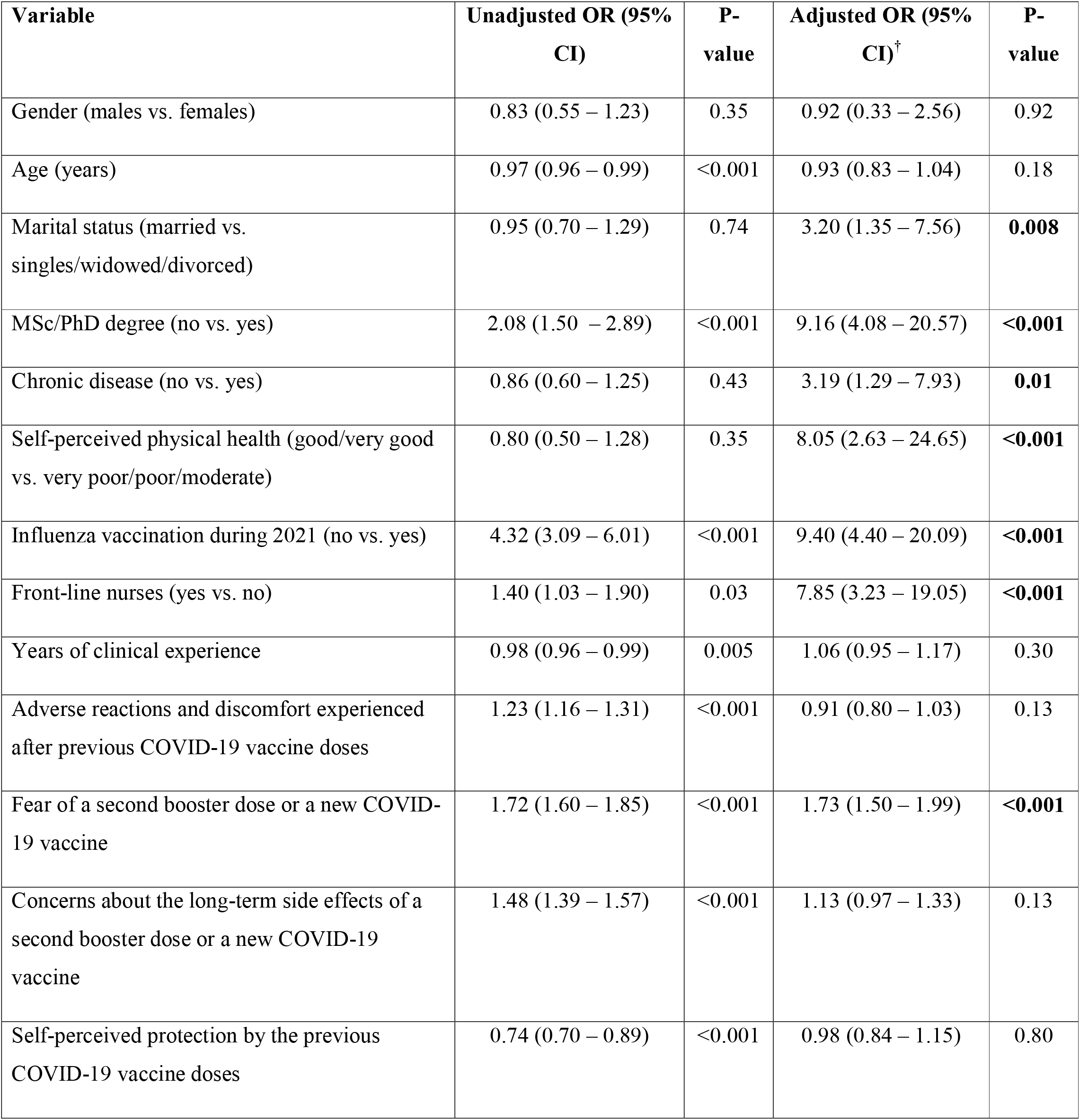
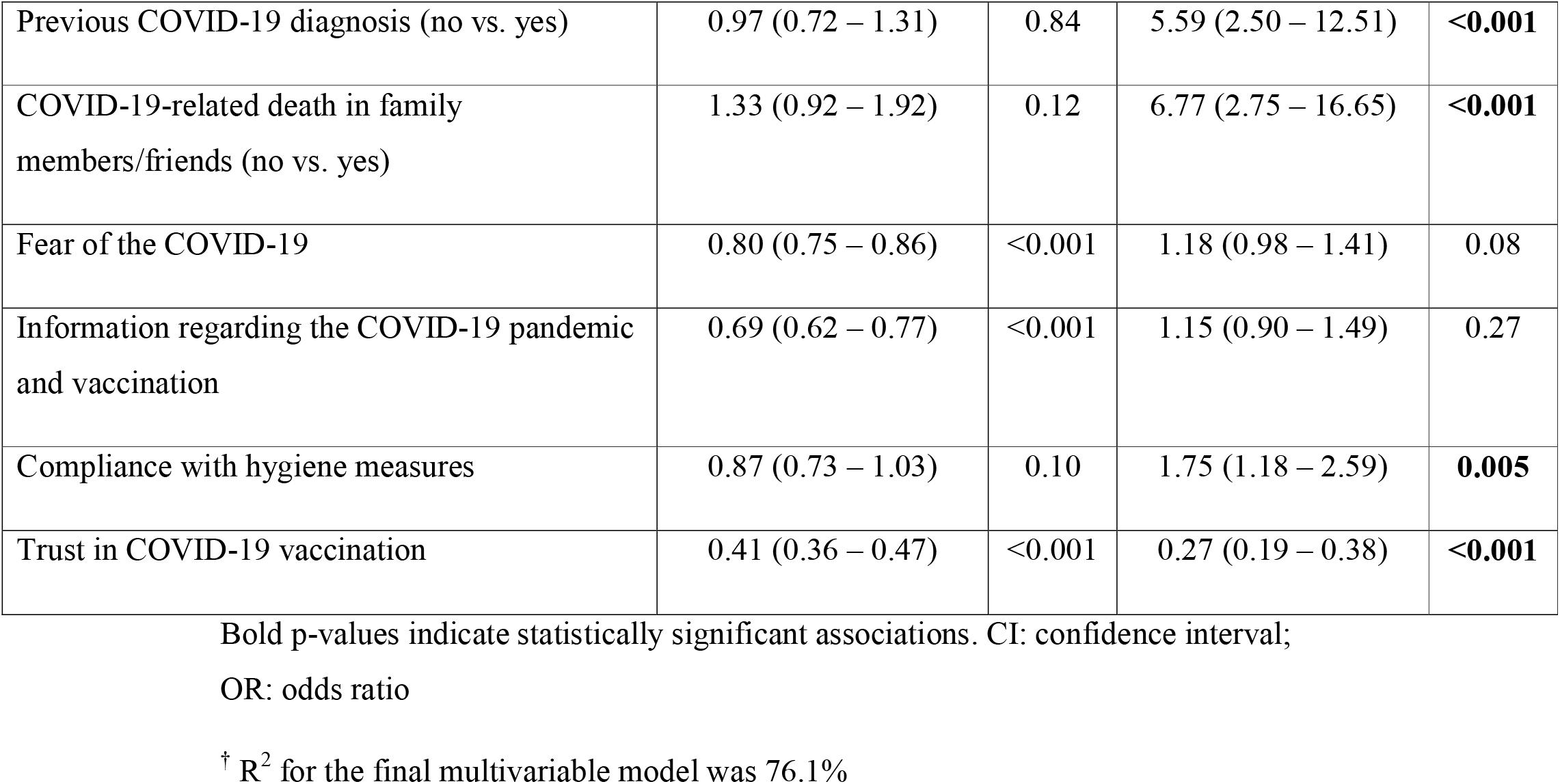
Univariate and multivariable logistic regression analysis with second booster dose/new COVID-19 vaccine hesitancy as the dependent variable (reference: willing participants to accept vaccination).

| Variable | Unadjusted OR (95% CI) | P-value | Adjusted OR (95% CI)<sup>†</sup> | P-value |
| --- | --- | --- | --- | --- |
| Gender (males vs. females) | 0.83 (0.55 – 1.23) | 0.35 | 0.92 (0.33 – 2.56) | 0.92 |
| Age (years) | 0.97 (0.96 – 0.99) | <0.001 | 0.93 (0.83 – 1.04) | 0.18 |
| Marital status (married vs. singles/widowed/divorced) | 0.95 (0.70 – 1.29) | 0.74 | 3.20 (1.35 – 7.56) | **0.008** |
| MSc/PhD degree (no vs. yes) | 2.08 (1.50 – 2.89) | <0.001 | 9.16 (4.08 – 20.57) | **<0.001** |
| Chronic disease (no vs. yes) | 0.86 (0.60 – 1.25) | 0.43 | 3.19 (1.29 – 7.93) | **0.01** |
| Self-perceived physical health (good/very good vs. very poor/poor/moderate) | 0.80 (0.50 – 1.28) | 0.35 | 8.05 (2.63 – 24.65) | **<0.001** |
| Influenza vaccination during 2021 (no vs. yes) | 4.32 (3.09 – 6.01) | <0.001 | 9.40 (4.40 – 20.09) | **<0.001** |
| Front-line nurses (yes vs. no) | 1.40 (1.03 – 1.90) | 0.03 | 7.85 (3.23 – 19.05) | **<0.001** |
| Years of clinical experience | 0.98 (0.96 – 0.99) | 0.005 | 1.06 (0.95 – 1.17) | 0.30 |
| Adverse reactions and discomfort experienced after previous COVID-19 vaccine doses | 1.23 (1.16 – 1.31) | <0.001 | 0.91 (0.80 – 1.03) | 0.13 |
| Fear of a second booster dose or a new COVID-19 vaccine | 1.72 (1.60 – 1.85) | <0.001 | 1.73 (1.50 – 1.99) | **<0.001** |
| Concerns about the long-term side effects of a second booster dose or a new COVID-19 vaccine | 1.48 (1.39 – 1.57) | <0.001 | 1.13 (0.97 – 1.33) | 0.13 |
| Self-perceived protection by the previous COVID-19 vaccine doses | 0.74 (0.70 – 0.89) | <0.001 | 0.98 (0.84 – 1.15) | 0.80 |
| Previous COVID-19 diagnosis (no vs. yes) | 0.97 (0.72 – 1.31) | 0.84 | 5.59 (2.50 – 12.51) | **<0.001** |
| COVID-19-related death in family members/friends (no vs. yes) | 1.33 (0.92 – 1.92) | 0.12 | 6.77 (2.75 – 16.65) | **<0.001** |
| Fear of the COVID-19 | 0.80 (0.75 – 0.86) | <0.001 | 1.18 (0.98 – 1.41) | 0.08 |
| Information regarding the COVID-19 pandemic and vaccination | 0.69 (0.62 – 0.77) | <0.001 | 1.15 (0.90 – 1.49) | 0.27 |
| Compliance with hygiene measures | 0.87 (0.73 – 1.03) | 0.10 | 1.75 (1.18 – 2.59) | **0.005** |
| Trust in COVID-19 vaccination | 0.41 (0.36 – 0.47) | <0.001 | 0.27 (0.19 – 0.38) | **<0.001** |

Bold p-values indicate statistically significant associations. CI: confidence interval;
OR: odds ratio
<sup>†</sup> R<sup>2</sup> for the final multivariable model was 76.1%

## 5. Discussion

To the best of our knowledge, this is the first study that evaluates the second booster dose/new COVID-19 vaccine hesitancy in a sample of fully vaccinated nurses. Moreover, we investigated the potential predictor variables of vaccine hesitancy among the study population. Our study was conducted after the approval of a second booster dose only for vulnerable fully vaccinated groups, by the Greek Committee on Vaccination. Thus, at the time of the study, a second COVID-19 booster dose was not recommended to healthcare workers in Greece.

We found that a significant percentage of nurses (30.9%) were hesitant toward a second booster dose or a new COVID-19 vaccine. Since literature suggests a predictable relationship between the intentions of healthcare workers and their subsequent behavior (Eccles et al., 2006), high levels of nurses’ vaccine hesitancy toward a second booster dose/new COVID-19 vaccine could be a significant threat for healthcare systems over the winter period. Studies in the USA, Czech, Saudi Arabia, and Singapore estimated healthcare workers hesitancy toward the first booster dose and found a hesitancy percentage of 33%, 28.8%, 44.7%, and 26.2% respectively (Alhasan et al., 2021; Chrissian et al., 2022; Klugar et al., 2021; Koh et al., 2022). The situation is even worse if we consider that the median value of hesitancy toward the first booster is higher among healthcare workers compared to the general population (31% vs. 22.1%) (Al-Qerem et al., 2022; Chu et al., 2022; Lai et al., 2021; Lounis et al., 2022; Miao et al., 2022; Paul & Fancourt, 2022; Rzymski et al., 2021; Wu et al., 2022; Yoshida et al., 2022). High levels of hesitancy toward a second booster dose/new COVID-19 vaccine in our study should be an alarm bell to the policymakers to scale up effective COVID-19 vaccination programmes in order to disseminate reliable information about the various highly contagious variants of SARS-CoV-2 and provide recommendations to healthcare workers about receiving an additional vaccine dose. It would be crucial to achieve a high acceptance rate of a second booster dose/new COVID-19 vaccine among nurses since they are the front-line healthcare workers during the COVID-19 pandemic. Also, high levels of vaccine hesitancy among nurses could be a reason for even higher hesitancy among the general population since nurses are often viewed as role models of health behaviors (Blake & Harrison, 2013).

The results presented in this study suggest that the primary reasons for refusing a second booster/new COVID-19 vaccine were concerns about side effects, safety and effectiveness of an additional dose. These findings are confirmed by a study in the USA that investigated the attitudes of vaccinated healthcare workers toward the first booster dose (Chrissian et al., 2022). Individuals’ considerations regarding side effects, safety and effectiveness of booster doses are also confirmed by studies in the general population (Al-Qerem et al., 2022; Lai et al., 2021). Moreover, it is well known that vaccine safety concerns are a significant source of initial COVID-19 vaccine hesitancy among healthcare workers (Galanis et al., 2021a; Li et al., 2021). In addition, a systematic review in the first year of the pandemic revealed that nurses had higher COVID-19 vaccine hesitancy due to concerns regarding safety and efficacy (Al□Amer et al., 2022).

We found that decreased trust in COVID-19 vaccination was associated with increased hesitancy. This finding is confirmed by a national cross-sectional study in Saudi Arabia investigating first vaccine booster acceptability among healthcare workers (Alhasan et al., 2021). Moreover, several studies suggest that trust in COVID-19 booster doses increases willingness of the public to accept a first COVID-19 booster (Lai et al., 2021; Lounis et al., 2022; Rzymski et al., 2021). Low levels of confidence in COVID-19 are a significant obstacle for booster doses uptake and policymakers should strength public trust in booster safety. Also, COVID-19 vaccination programmes are critical for the control of the pandemic worldwide and these efforts cannot succeed without public confidence. Thus, implementation of a post-approval vaccine safety monitoring system of the COVID-19 booster doses in the real world can provide an essential safety net (Dhanda et al., 2022; Lee et al., 2020). Post-licensure observational studies for safety and effectiveness of COVID-19 booster doses are necessary in order to provide reliable information and enable timely and accurate policy decisions.

Our results showed that increased fear of a second booster dose or a new COVID-19 vaccine was associated with increased hesitancy. Side effects after COVID-19 vaccination are common, but they are usually mild and self-limited (Dighriri et al., 2022). Moreover, literature suggest that side effects after the primer COVID-19 vaccine doses decreases intention of the individuals to accept the first booster dose (Al-Qerem et al., 2022; Lounis et al., 2022; Miao et al., 2022; Rzymski et al., 2021; Wu et al., 2022). Chrissian et al. found that healthcare workers who missed work due to vaccine-related symptoms after the primer doses were less likely to accept future booster doses (Chrissian et al., 2022). Therefore, it is reasonable for healthcare workers to fear another booster dose especially if they have already experienced side effects after previous COVID-19 vaccinations. Since consequences of side effects may impact future booster doses acceptance, more efforts should be made including dissemination of information from valid sources.

In our study, healthcare workers who did not receive a flu vaccine the previous season were less likely to accept a second COVID-19 booster dose or a new COVID-19 vaccine. A systematic review confirms this finding since influenza vaccination among healthcare workers is associated with primer COVID-19 vaccine doses acceptance (Galanis et al., 2021b). In addition, healthcare workers with positive attitudes towards COVID-19 vaccination are more likely to accept a COVID-19 vaccine (Galanis et al., 2021b). Unfortunately, vaccine hesitancy among healthcare workers continues to be a major public health issue (Lau et al., 2020; Wilson et al., 2020). Thus, the hesitancy of nurses to accept future COVID-19 booster doses could diminish the public trust (Opel et al., 2013). Nurses should act as role models in order to build confidence in the public and to motivate individuals to adopt health behaviours such as vaccination.

It is notable that in our study population, nurses without a chronic disease and those with good/very good physical health were less likely to accept a second COVID-19 booster dose/new COVID-19 vaccine. It seems that healthy nurses are less willing to get vaccinated against COVID-19. Literature confirms our findings since healthcare workers with chronic conditions are more willing to accept the primer COVID-19 vaccine doses (Wang et al., 2020). Moreover, public acceptance of the first booster dose is higher among individuals with chronic diseases (Lounis et al., 2022; Paul & Fancourt, 2022; Rzymski et al., 2021). Self-perceived risk of infection is associated with vaccine acceptance (Gagneux-Brunon et al., 2021), while healthcare workers considering themselves at risk of severe COVID-19 are more likely to accept a COVID-19 vaccine (Neumann-Böhme et al., 2020). Nurses who have chronic conditions and are in bad health should be identified as a high-risk group by policymakers to accept a second COVID-19 booster dose/new COVID-19 vaccine when it is recommended.

Our study showed that increased compliance with hygiene measures was associated with increased vaccine hesitancy. Probably this finding could be attributed to the fear of COVID-19. In particular, there is a relation between compliance and fear since compliance with recommendations is associated with high fear of COVID-19 (Håkansson & Claesdotter, 2022). Moreover, as the level of fear of COVID-19 increases, willingness of individuals to accept a COVID-19 vaccine increases (Mertens et al., 2022; Patelarou et al., 2022; Seddig et al., 2022). There is a need for policymakers to address COVID-19 vaccination hesitancy by emphasizing factors that support acceptance of vaccines, such as decrease of fear of COVID-19.

We found that booster hesitancy was higher among nurses without a MSc/PhD degree. Several studies in both healthcare workers and the general population confirm the fact that lower educational level is associated with decreased acceptance of the first COVID-19 booster dose (Chrissian et al., 2022; Chu et al., 2022; Miao et al., 2022; Paul & Fancourt, 2022). This could be attributed to the fact that educational level is associated with health knowledge regarding COVID-19 (Gomes da Silva et al., 2021). Increased knowledge regarding COVID-19 predicts first booster willingness in healthcare workers (Alhasan et al., 2021). Valid, efficient, effective, and continuous information regarding COVID-19 vaccination should be provided to nurses through appropriate sources to increase understanding about safety, efficacy, and effectiveness of COVID-19 vaccines.

### 5.1. Limitations

Our study had several limitations. We conducted an on-line cross-sectional study and convenience sample was unavoidable. A random sampling technique would be more ideal but it was not feasible since we cannot have access to all the nurses in the Greece. Additionally, as the number of nurses who received the questionnaire was not known, we cannot calculate the response rate and we cannot find possible differences between participants and non-participants. Moreover, our study suffers from selection bias since it is expected that participation rate would be higher among nurses who are interested in health issues, e.g. COVID-19 vaccination. Therefore, generalization of our results is not possible. We investigated several predictors of second COVID-19 booster dose/new COVID-19 vaccine hesitancy among nurses but there is still room in this research topic. For instance, further studies could explore income, healthcare facilities (hospitals, health centers, nursing homes, etc.), knowledge, sources of information, and psychosocial factors as possible predictors of vaccine hesitancy. In addition, our cross-sectional study just reflects a snapshot of nurses’ hesitancy to accept future COVID-19 vaccine doses. Thus, further studies in the future should investigate nurses’ intention since attitudes can change over time. Finally, we used a self-administrated questionnaire and self-reporting bias is probable. In that case, hesitancy rate could be an underestimation due to social desirability.

## 6. Conclusions

Our study revealed that a significant percentage of nurses were hesitant toward a second booster dose/new COVID-19 vaccine. Also, our data present important insights on factors that relate to COVID-19 vaccine hesitancy providing guidance to policymakers on the implementation of future COVID-19 vaccination programmes. Nurses’ initial hesitancy could be a barrier to efforts to control the COVID-19 pandemic especially during the winter period. Moreover, new highly contagious variants of SARS-CoV-2 are still a possible scenario, while evidence shows that existing COVID-19 vaccines offer protection only for limited time periods. This is bad news and additional COVID-19 vaccine doses seem to be the best solution.

## 7. Relevance to clinical practice

Nurses’ role during the COVID-19 pandemic is essential since they are the front-line healthcare workers empowering the public with their passion and empathy. Therefore, fully vaccinated nurses against COVID-19 will likely play an important role in controlling the pandemic through vaccines. There is a need to communicate COVID-19 vaccine science in a way that is accessible to nurses in order to decrease COVID-19 vaccine hesitancy.

### What does this paper contribute to the wider global clinical community?

- We found that a significant percentage of nurses were hesitant toward a second booster dose/new COVID-19 vaccine.
- Vaccine hesitancy among nurses is crucial since nurses can act as role models for the public.
- High acceptance rate of a second booster dose/new COVID-19 vaccine among nurses is necessary to control the pandemic, over the winter period.

## Data Availability

All data produced in the present study are available upon reasonable request to the authors

## Acknowledgments

None

